# Healthy Lifestyles and Post-Stroke Depression: A NHANES-Based Cross-sectional Study of Stroke Survivors

**DOI:** 10.1101/2025.04.14.25325843

**Authors:** Chenyiyi He, Xuehui Fan, Li Gao, Guojun Yun, Zhijiang Tan, Yucun Chen, Li Li, Jianguo Cao

## Abstract

**Background:** Post-stroke depression (PSD) is a common complication that seriously affects the recovery and quality of life of stroke survivors. Although individual lifestyle factors are associated with reduced risk of depression, the combined associations remain unclear. The aim of this research was to investigate the relationship between combined healthy lifestyles and the risk of PSD.

**Methods:** We analyzed data from the National Health and Nutrition Examination Survey (NHANES) 2005–2020, including 715 stroke survivors (mean age: 61.3 years, 51.61% male). A healthy lifestyle score (0–5) was constructed based on adherence to five favorable lifestyle behaviors, including current nonsmoking, low-to-moderate alcohol drinking, adequate physical activity, healthy diet, and optimal waist circumference. PSD was assessed using the Patient Health Questionnaire-9 (PHQ-9), with a score ≥9 indicating depression. Multivariable logistic regression models, adjusted for demographic and clinical confounders, were used to estimate associations.

**Results:** In this study population, 68.53% were non-smokers, 55.10% drank alcohol moderately, 18.04% exercised regularly, 40.00% ate a healthy diet, and 13.15% had an ideal waist circumference. Compared to those with 0-1 health behaviors, those with 3-5 had a 52% lower risk of PSD (OR=0.48, 95% CI: 0.29-0.78); each additional health behavior was associated with a 30% lower risk of PSD (OR=0.70, 95% CI. 0.58-0.84). This result persisted across sensitivity analyses.

**Conclusion:** Adherence to healthy lifestyles may help reduce the risk of PSD. This result highlights the crucial role of incorporating lifestyle-based interventions into post-stroke rehabilitation, and the potential benefits to both physical and mental health.

## Introduction

Globally, stroke is a leading cause of death and disability, often due to a blocked or ruptured brain vessel, damaging neurological function and quality of life^[1]^.It is also estimated that 15 million people have a stroke each year, with one-third of them dead from it, and another one-third permanently disabled^[2]^. The global burden of stroke continues to be rising, notably in low- and middle-income countries, as influenced by aging populations, sedentary lifestyles, unhealthy diets, and the increased prevalence of chronic diseases ^[3]^. Post-stroke depression (PSD) is a common neuropsychiatric outcome following stroke, often impairing cognitive processes, hindering functional recovery, and significantly reducing patients’ quality of life.^[4]^. Evidence suggests that between 40% and 50% of individuals who have experienced a stroke develop depressive symptoms within the first year post-event.^[5]^.Stroke survivors suffering from PSD face a 50% higher mortality risk, a 1.49-fold increase in recurrence rates, and tend to endure prolonged disability compared to those without depression.^[6]^,^[7]^, ^[8]^As a corollary, PSD not only hinders functional recovery, but also increases the burden on on families and the healthcare system^[9]^. Therefore, identifying effective strategies to mitigate PSD risk is essential for improving both physical and mental health outcomes in stroke survivors.

Mounting evidence suggested that lifestyle modifications, such as regular exercise, smoking cessation, a good balanced diet and moderate alcohol consumption, are vital in reducing the risk of chronic disease and improving mental health^[10]^,^[11]^. Studies have shown that sticking to a healthy diet, regular exercise and avoiding smoking significantly reduces the risk of cardiovascular disease and metabolic disorders, both of which are closely linked to the occurrence of stroke^[12]^,^[13]^. Regular exercise is particularly important in stroke prevention^[14]^. In addition, stroke survivors with higher health literacy are more likely to adopt positive health behaviors, such as regular exercise and a balanced diet, that promote mental health^[15]^. Social support, adequate nutrition and physical activity are crucial in decreasing the prevalence of depression, especially for vulnerable populations^[16]^.

Despite the well-established positive impact of a healthy lifestyle on mental health, existing research primarily focuses on the role of individual factors in stroke prevention and rehabilitation.There has been relatively limited research on the combined effects of multiple lifestyle factors, especially on post-stroke depression (PSD). In addition, some studies have revealed that a healthy lifestyle may help decrease the risk of depression, but these conclusions are mostly based on the general population or patients with chronic diseases.It is still lacking systematic exploration of how these factors synergistically affect PSD in stroke survivors. This lack of comprehensive evidence hampers our understanding of the underlying mechanisms of PSD and restricts the refinement of effective rehabilitation approaches.

In response to this research gap, we analyzed data from the nationally representative NHANES survey, aiming to explore how lifestyle behaviors collectively relate to post-stroke depression. We aim to provide a scientific basis and practical guidance for more effective PSD intervention and management strategies through a comprehensive analysis of these behaviors.

## Methods

### Study population

From the NHANES survey cycles spanning 2005 to 2020, we identified 44,728 non-pregnant adults, aged between 20 and 79 years, for eligibility evaluation. A total of 770 stroke survivors were left after excluding 20,873 subjects with missing lifestyle information and 23,085 subjects without history of stroke. In addition, 55 missing covariate subjects were excluded. The count and percentage of missing covariates are displayed in Table S1, and the eligibility screening process is shown in Figure S1.

### Construction of healthy lifestyle score

A healthy lifestyle score was constructed by aggregating five modifiable lifestyle factors: non-smoking status, moderate alcohol intake, adequate physical activity, adherence to a balanced dietary pattern, and maintenance of an optimal waist circumference^[17]^. Each factor was assigned a binary score (0 = unhealthy, 1 = healthy), with total scores ranging from 0 to 5.

Higher scores indicate progressively greater adherence to health-promoting behaviors, where a score of 5 represents full compliance with all recommended lifestyle criteria. Specific definitions and thresholds for each health behavior are provided in Supplementary Table S2.

### Definitions of depression

Depressive symptoms were assessed using the Patient Health Questionnaire-9 (PHQ-9), a validated self-report instrument designed to evaluate the frequency of depression-related symptoms over a couple of weeks. The PHQ-9 comprises nine items scored on a 0–3 Likert scale (0 = “not at all” to 3 = “nearly every day”), yielding a total score up to 27. A cutoff score of ≥9 was employed to define clinically significant depression ^[18][19]^.

### Assessment of covariates

In this study, we collected covariates on demographic characteristics, lifestyle factors, and socioeconomic status. The variables included race/ethnicity, marital status, education level, income status (assessed via the poverty income ratio [PIR]), blood pressure, and metabolic health indicators relevant to hypertension and diabetes.

### Statistical methods

We conducted a weighted multivariate logistic regression analysis to examine the association between healthy lifestyle scores—reflecting factors such as diet, physical activity, and smoking habits—and the risk of depression among stroke survivors. Multicollinearity was evaluated using variance inflation factors (VIFs), and as reported in Table S3, all VIFs were substantially below the threshold of 10, confirming the absence of significant multicollinearity. In multivariable model 1, adjustments were made for fundamental clinical characteristics, including age, sex, and race/ethnicity. In multivariable model 2, we further adjusted for additional covariates, namely marital status, family poverty, education status, and comorbidities.

To ensure the robustness of our findings, we performed stratified and interaction analyses across various subgroups. Interaction effects were tested using likelihood ratio tests, and the relative excess risk due to interaction (RERI) was calculated to assess additive interactions, as detailed previously ^[20]^. To further validate our results, we conducted a series of sensitivity analyses: we redefined the criteria for healthy drinking, adjusted for confounding using propensity scores, applied multiple imputation to address missing data, and calculated E-values to evaluate the potential impact of residual confounding. Detailed methodologies for these analyses are available in the supplementary section.

For healthy participants, continuous variables were compared using analysis of variance (ANOVA) or the Student’s t-test, as appropriate, while categorical variables were analyzed with the chi-square test or Fisher’s exact test, depending on sample size and expected frequencies. Statistical significance was defined as a two-sided P-value <0.05.

## Results

### Baseline Characteristics

Overall, 715 stroke survivors (mean age 61.30 years and 51.61% male) were included in the current study (Table 1).Individuals with a greater number of healthy lifestyle factors exhibited associations with being of advanced age, male sex, married status, higher educational attainment, superior socioeconomic status, elevated Healthy Eating Index-2015 (HEI-2015) scores, and reduced body mass index (BMI) and waist circumference measurements (all P < 0.05; Table S4).

**Table 1.** Characteristics of stroke survivors included in the study.

| **Characteristics<sup>a</sup>** | **Overall (N=715)** |
| --- | --- |
| Age, years | 61.30 (11.75) |
| BMI, kg/m<sup>2</sup> | 31.13 (7.37) |
| Waist circumference, cm | 106.54 (16.49) |
| HEI-2015 | 48.59 (11.23) |
| Male, n (%) | 369 (51.61) |
| Race/ethnicity, n (%) |  |
| Non-Hispanic white | 325 (45.45) |
| Non-Hispanic black | 245 (34.27) |
| Mexican American | 60 (8.39) |
| Others | 85 (11.89) |
| Marital status, n (%) |  |
| Married | 352 (49.23) |
| Others | 363 (50.77) |
| Education attainment, n (%) |  |
| Under high school | 194 (27.13) |
| High school | 217 (30.35) |
| Above high school | 304 (42.52) |
| Family PIR, n (%) |  |
| <1.3 | 301 (42.10) |
| 1.3-<3.5 | 282 (39.44) |
| ≥3.5 | 132 (18.46) |
| Current nonsmoking, n (%) | 490 (68.53) |
| Low-to-moderate alcohol drinking, n (%) | 394 (55.10) |
| Adequate physical activity, n (%) | 129 (18.04) |
| Healthy diet, n (%) | 286 (40.00) |
| Optimal waist circumference, n (%) | 94 (13.15) |
| No. of healthy lifestyle factors, n (%) |  |
| 0 | 51 (7.13) |
| 1 | 218 (30.49) |
| 2 | 227 (31.75) |
| 3 | 162 (22.66) |
| 4 | 50 (6.99) |
| 5 | 7 (0.98) |
| Hypertension, n (%) | 581 (81.26) |
| Diabetes, n (%) | 273 (38.18) |

<sup>a</sup> Continuous variables were expressed as mean (SD), and categorical variables were presented as number (percentage).
Abbreviations: BMI, body mass index; HEI, Healthy Eating Index; PIR, poverty-income ratio.

### Healthy Lifestyle Score and Depression Risk Association

In unadjusted analyses, stroke survivors with 3–5 healthy lifestyle factors demonstrated a 66% reduction in depression risk (odds ratio [OR]: 0.34; 95% confidence interval [CI]: 0.21–0.53) compared to those with 0–1 scores (Table 2). After full adjustment for potential confounders, adherence to 3–5 healthy lifestyle factors was associated with a 52% descreased risk of depression (OR: 0.48; 95% CI: 0.29–0.78). Furthermore, each additional healthy lifestyle factor was linked to a 30% reduction in depression risk (OR: 0.70; 95% CI: 0.58–0.84), indicating a dose-dependent protective effect (Table 2). Stratified analyses revealed no significant multiplicative interactions between healthy lifestyle factors and demographic variables (all P-interaction >0.05). However, additive-scale interaction analysis identified a statistically significant synergistic effect between age and combined healthy lifestyle factors on depression risk reduction (RERI: 0.32; 95% CI: 0.03–0.60) ( Table S6).

**Table 2.** Association of healthy lifestyle score with risk of depression among stroke survivors.

| Variable | No. of healthy lifestyle factors |  |  | Each additional healthy lifestyle factor |
| --- | --- | --- | --- | --- |
|  | 0-1 | 2 | 3-5 |  |
| Case/total (%) | 84/269<br>(31.23) | 48/227<br>(21.15) | 29/219<br>(13.24) |  |
| Crude model | 1.00<br>(reference) | 0.59 (0.39-0.89) | 0.34 (0.21-0.53) | 0.61 (0.51-0.73) |
| Model 1<sup>a</sup> | 1.00<br>(reference) | 0.64 (0.42-0.98) | 0.39 (0.24-0.63) | 0.65 (0.54-0.78) |
| Model 2<sup>b</sup> | 1.00<br>(reference) | 0.69 (0.45-1.05) | 0.48 (0.29-0.78) | 0.70 (0.58-0.84) |

<sup>a</sup> Model 1 was adjusted for age (<60, ≥60 years), sex (male, female), and race/ethnicity (non-Hispanic white, others).
<sup>b</sup> Model 2 was further adjusted for marital status (married, others), family poverty-income ratio (<3.5, ≥3.5), education attainment (above high school, high school and below), hypertension (yes, no), and diabetes (yes, no).

Analysis of individual lifestyle factors revealed that current nonsmoking (OR: 0.66, 95% CI: 0.44–1.00) and adequate physical activity (OR: 0.43, 95% CI: 0.22–0.78) were significantly associated with reduced depression risk (Table S7). Low-to-moderate alcohol consumption, healthy dietary patterns, and optimal waist circumference showed non-significant trends toward lower depression risk. Table S8 indicated that the protective association between a composite healthy lifestyle score and depression was attenuated when excluding current nonsmoking and adequate physical activity. Specifically, the ORs for depression (comparing 3-4 vs. 0-1 healthy lifestyle factors) increased upon exclusion of these two factors, suggesting their substantial contribution to the observed protective effect.

### Sensitivity analyses

The robustness of our results was evlauted the following sensitivity analyses ( Tables S9-S14). First, we redefined moderate alcohol consumption to include non-drinkers, which did not significantly alter the results (Table S9). Second, after applying propensity score (PS) adjustment to account for potential confounders, the association remained significant (OR: 0.47, 95% CI: 0.29–0.76, Table S10). Third, after multiple imputation (MI), the results remained consistent ( Table S11). A 3.59 E-value suggested that it would take very strong confounding to negate the inverse association observed in our study ( Table S12). Finally, weighted healthy lifestyle score model showed that adequate physical activity contributed most (β=0.46), followed by current nonsmoking (β=0.23), healthy diet (β=0.16), low-to-moderate drinking (weighted β=0.10), and optimal waist circumference (β=0.05) ( Table S13). In full-adjusted model, stroke survivors with the highest weighted healthy lifestyle score tertile were confronted with 57% (OR: 0.43, 95% CI: 0.26-0.69) decreased risk of depression as compared with the lowest weighted healthy lifestyle score tertile (see Supplementary Table S14). To visualize the dose-response relationship, we performed restricted cubic spline (RCS) analysis, which confirmed a linear inverse association between the weighted lifestyle score and PSD risk (P-overall < 0.001, P-nonlinearity = 0.437, Figure 1).

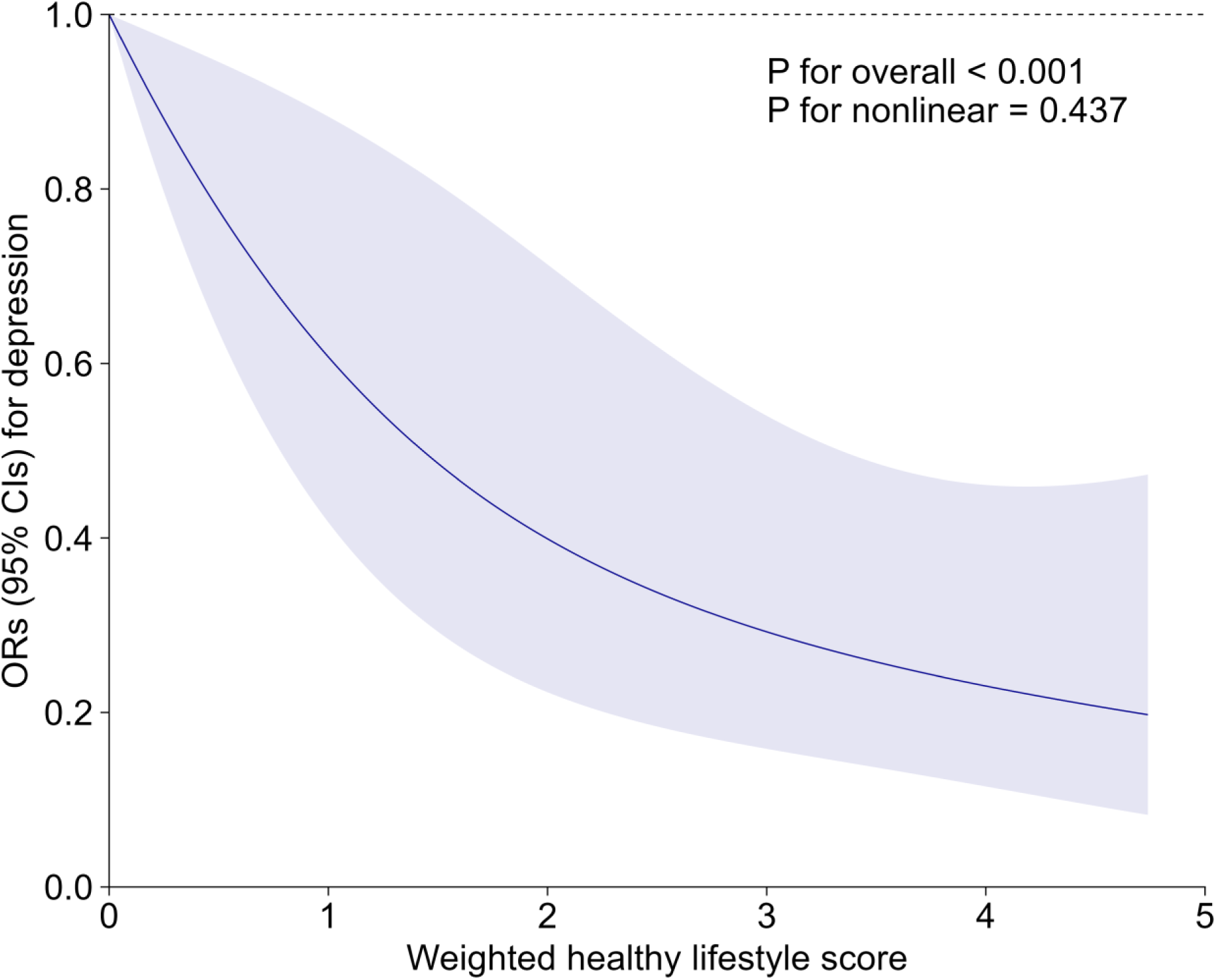

## Discussion

In this nationally representative cross-sectional study, adopting combined healthy lifestyle factors was associated with a lower risk of depression among stroke survivors. This inverse relationship remained robust in sensitivity analyses. Notably, when current nonsmoking and adequate physical activity were removed from the lifestyle score, the association was no longer evident, suggesting individual lifestyle factors may differ in their contributions.

Our study identified physical activity as an important protective factor in reducing the risk of post-stroke depression (PSD), a finding that is consistent with previous studies. For example, a meta-analysis by Li et al. ^[21]^ that included 36 studies involving 1,477 middle-aged and elderly stroke patients showed that exercise training can significantly alleviate depressive symptoms. Similarly, a prospective observational study by Thilarajah et al. ^[22]^ found that patients with a high level of physical activity before stroke had fewer depressive symptoms three months after stroke. These findings further support the protective effect of physical activity on PSD observed in our study and suggest that incorporating exercise interventions into stroke rehabilitation programs may effectively reduce depressive risk and enhance patients’ quality of life.

This study observed a association between smoking and increased risk of post-stroke depression (PSD), which is consistent with multiple prior studies. For instance, one study examining predictors of PSD among patients with minor ischemic stroke reported a significant relationship between smoking and the occurrence of PSD^[23]^. Additionally, a prospective study by Shi et al. ^[24]^, involving 5,000 patients, further confirmed smoking as an independent risk factor for PSD. Nevertheless, Tennen et al.^[25]^ ^[25]^ found no significant association between smoking and PSD but suggested that hypertension may have a greater impact on post-stroke emotional outcomes. This finding should be interpreted with caution due to small sample size. These results highlight a link between smoking and an increased risk of PSD.

Beyond regular exercise and giving up smoking, maintaining a balanced diet and proper weight management also play key roles in reducing the likelihood of post-stroke depression (PSD). Research has identified a notable link between unhealthy eating habits—especially those heavy in sugar and fat—and an increased chance of developing depression ^[26]^,^[27]^. Surprisingly, even among relatively healthy populations, diet plays a decisive role in mental well-being. The Whitehall II cohort study, which tracked 3,486 middle-aged individuals, found that adherence to a “whole-food” diet rich in vegetables, fruits, and fish was linked to a lower risk of depression, whereas high consumption of processed foods significantly increased this risk ^[28]^. Moreover, a well-balanced diet has been shown not only to alleviate depressive symptoms but also to slow cognitive decline following stroke ^[29]^.Given the profound impact of diet on psychological and cognitive health, incorporating dietary interventions into stroke rehabilitation programs may offer a novel approach to reducing the risk of PSD.

Additionally, the relationship between alcohol consumption and post-stroke depression (PSD) warrants attention. A national cohort study among US adults found that alcohol use disorders increased depression risk, whereas depression did not conversely increase alcohol use disorder risk ^[30]^. Evidence suggests that excessive alcohol intake may disrupt neurotransmitter function, elevate anxiety and depressive symptoms, and interact with other unhealthy lifestyle behaviors (e.g., smoking, physical inactivity), exacerbating the psychological burden in stroke patients^[31]^. Therefore, appropriate management of alcohol intake should be emphasized in PSD prevention and treatment to reduce adverse psychological effects and improve stroke recovery outcomes.

Growing evidence indicates that lifestyle factors—such as diet, physical activity, smoking, and alcohol consumption—significantly influence post-stroke depression (PSD). However, the underlying mechanisms remain poorly understood. The pathophysiology of PSD is complex, involving multiple interrelated biological processes, such as hypothalamic-pituitary-adrenal (HPA) axis dysregulation^[32]^, neuroinflammation ^[33]^, neurotransmitter imbalances^[34]^, and oxidative stress ^[35]^. These mechanisms collectively drive the onset and progression of PSD. After a stroke, HPA axis dysregulation and increased oxidative stress may induce neuroinflammation, elevating pro-inflammatory cytokines such as IL-1, IL-6, and TNF-α^[36]^.

Neurotransmitter system disruptions concurrently reduce serotonin (5-HT), norepinephrine (NE), and dopamine (DA) levels, exacerbating PSD symptoms ^[37]^. Lifestyle factors may influence these pathological processes and thus affect PSD risk. Regular physical activity has been shown to promote the expression of BDNF, which plays a vital role in neural repair and synaptic plasticity. It also helps to suppress inflammatory pathways and support neurotransmitter regulation, ultimately reducing the risk of post-stroke depression ^[38]^. Similarly, maintaining a balanced diet may benefit neurological health by modulating the gut-brain axis and lowering oxidative stress, which in turn supports mood stability and cognitive function^[39]^.

Conversely, unhealthy lifestyle choices may heighten PSD susceptibility. Diets rich in saturated fat and refined sugar, along with tobacco use, have been implicated in exacerbating neuroinflammatory responses and oxidative damage—both key contributors to the development of post-stroke depression **^Error! Reference source not found.^**.Moreover, Long-term alcohol use can interfere with the BDNF–serotonin interaction in the brain, potentially impairing neuroplasticity and emotional regulation, which may increase vulnerability to PSD^[40]^ While direct evidence is limited, alcohol’s neurotoxicity may worsen neural dysfunction and mood disturbances, increasing PSD risk.

Although this study’s findings align with most prior research, some discrepancies persist. For example, Delhey et al^[41]^. ^[41]^conducted a meta-analysis suggesting that diet and exercise had limited effects on depression, whereas socioeconomic factors might be more influential. However, the small sample size (N=121) may have limited its statistical power and generalizability.

This study drew upon a nationally representative dataset, thereby enhancing the generalizability of its results. Nevertheless, certain limitations should be highlighted. First, due to its observational design, the study cannot ascertain causation. Further longitudinal research or randomized controlled trials (RCTs) were required for establishing causal relationships. Second, relying on self-reported lifestyle measures may introduce recall bias and affect data accuracy.

Third, although multiple confounders were accounted for, the possibility of unmeasured residual confounding cannot be entirely ruled out. However, E-value analysis suggests that confounder of considerable strength could negate the observed inverse relationship. Finally, the constraints of NHANES did not permit an examination of long-term lifestyle alterations or their effects on PSD risk, emphasizing the need for further investigation in this domain.

Our study’s results suggested a association between multiple healthy lifestyle practices and a diminished likelihood of post-stroke depression. To apply these findings effectively in both public health and clinical contexts, integrating regular assessments and management protocols for depression among stroke survivors is recommended in community-based and primary care facilities. Meanwhile, multi-pronged education and intervention efforts that emphasize smoking cessation, limited alcohol consumption, healthy eating, and consistent physical activity can improve individuals’ awareness and adherence. Adopting a multidisciplinary framework enables thorough evaluations of each patient and supports the design of personalized strategies, such as exercise recommendations, dietary guidance, smoking-cessation programs, and psychological support, adjusted over time based on clinical progress and personal needs. Additionally, engaging family members and broader community resources can further reinforce healthy habits, thereby enhancing psychological recovery and overall quality of life for those affected by stroke.

### Conclusion

The present findings underscore the protective role of healthy lifestyle behaviors in mitigating the risk of post-stroke depression among individuals with a history of stroke.Notably, among the examined behaviors, maintaining regular physical activity and avoiding tobacco use appeared to offer the most substantial mental health benefits.Integrating multifaceted lifestyle strategies into post-stroke care may serve as a practical approach to enhancing both emotional recovery and general well-being.

## Data Availability

Data Availability Statement The data that support the findings of this study are publicly available from the National Health and Nutrition Examination Survey (NHANES) conducted by the Centers for Disease Control and Prevention (CDC). The NHANES datasets used in this analysis (2005?2020) are accessible through the CDC website at: https://www.cdc.gov/nchs/nhanes/index.htm. All data used in this study are de-identified and comply with relevant ethical guidelines.

